# COVID-19 Epidemic in Algeria: Assessment of the implemented preventive strategy

**DOI:** 10.1101/2020.04.21.20074443

**Authors:** Mohamed Hamidouche

## Abstract

Since the spread of the COVID-19 epidemic in China, several preventive strategies have been implemented worldwide to fight against the spread of SARS-CoV-2, in Algeria the control actions have been mainly social distancing, movement’s restriction, quarantine and lockdown.

In order to assess the preventive strategy adopted in Algeria, we studied three zones (whole country, the main outbreak at Blida, and whole country except Blida), we used as a indicator the basic reproduction number R0, we compared the mean R0-before and R0-after the implementation of the mitigation measures using bivariate analysis, then we used the results we found to estimate the number of COVID-19 cases avoided by these measures, then after, we predicted the peak of the epidemic’s first wave.

We note that the decrease in R0 after the preventive measures implementation was statistically significant (p value<10^−4^) in the three areas, thus, the preventive strategy in Algeria has been effective in its entirety. Our projection revealed that 15613 cases of COVID-19 were avoided in 14 days (from April 6, to April 19, 2020) in the whole country, and 1747 cases were prevented in Blida during the same period. We estimate that the peak of the first wave of the epidemic in Algeria will be reached with herd immunity of 15.93% as of November 2020, however, at least 56% of people with protective immunity would be needed to be sufficient to avoid a second wave. The used method to carry out the evaluation has given us a good overview, but, R0 is not the only factor to consider when estimating the burden of the epidemic, to do that, the number of daily admissions to intensive care units and hospitalizations remain good indicators.

In order to better the epidemic control in Algeria, it is possible to act on contact efficacy rate by adding an instruction to wear medical mask by ordinary population outside, this measure has been reported to be effective in some countries.

To conclude, it is good to keep in mind that a new way of life based on good hygiene habits and social distancing must therefore be in place and adopted by the population for at least several months, otherwise the demand for health care will exceed the availability.

## Introduction

In December 2019, consecutive pneumonia cases of unknown cause with clinical presentations resembling viral pneumonia appeared in Wuhan, Hubei, China, (Chaolin, et al., 2020), on December 31, 2019, the outbreak was reported for the first time at the World Health Organization, and investigations led to link it with Huanan seafood and animal market in Wuhan (WHO1, 2020). On January 7, 2020, the etiological agent of the outbreak was identified as a novel coronavirus, thus, it has been first named (2019-nCoV), then ‘severe acute respiratory syndrome coronavirus 2’ (SARS-CoV-2), while coronavirus disease associated with it is now referred to as Coronavirus Disease 2019 (COVID-19) since February 12, 2020. It is believed that the SARS-CoV-2 is animal-derived (WMHC, 2020), however, the source of the virus has not been well identified yet.

The reported data show that people of all ages are susceptible to SARS-CoV-2, however, links between COVID-19 and some risk factors like age and medical history have been established, so, elderly and people with underlying diseases tend to develop more severe forms once infected, also, obese people with BMI (Body Mass Index) of 40 or higher are at high-risk for severe illness from COVID-19 (CDC, 2020; Guan, et al., 2020; Chen, et al., 2020).

In Algeria, the first imported case of COVID-19 was reported on February 25, 2020, when an Italian national tested positive in Ouargla region in the south of the country, this case has been well isolated and only 37 cumulative cases are registered in this region until April 21, 2020. On March 1, 2020, a COVID-19 outbreak began in Blida region, northern Algeria, when two cases were reported after their contact with two Algerian nationals living in France, this region form a cluster of more than 5,4 million inhabitants with the surrounding cities (Algiers, Boumerdes, Tipaza) (Algerian Ministry of Health, 2020; ONSA, 2020).

Blida as a first outbreak and the origin of COVID-19 epidemic in Algeria remains the most affected area with cumulative cases number of 692 (24,6%) among a total of 2811 cases until April 21, 2020. The total case fatality rate was 13,94% (392 cases) as of April 21, 2020, distributed by age as follows; 2 case under 25 years old, 32 cases between 25 and 50 years old, 70 cases between 51 and 60 years old and 288 cases for people over 60 years old (Algerian Ministry of Health, 2020). A previous study has reported an average basic reproduction number R0=2.55 (95% CI 2.17-2.92) based on the actual incidence of the first 26 days, until March 19, 2020 (Hamidouche, 2020).

Algerian people in general and Blida inhabitants particularly are eager to know when they can resume their normal lives, so, this study aims to provide a scientific projection of COVID-19 epidemic’s parameters based on the implemented preventive strategy effectiveness (lockdown, self-isolation, social distancing, movement restrictions…etc.), to this end, we conduct a preventive strategy assessment in order to define the outbreak’s growth and to predict epidemic’s first wave peak, in addition to estimating the number of infections avoided, this will probably allow decision-makers to design their future plans to fight the COVID-19 epidemic in Algeria.

## Methods

In order to assess the implemented preventive strategy, we first observed and then performed a bivariate analysis of the average basic reproduction number (R0) before and after the implementation of control actions, after that, we established the divergence of COVID-19 cumulative cases numbers based on the found results, this has been done in three areas; i. Whole country, ii. Blida region, iii. Whole country except Blida.

### Data collection

We collected data on preventive and control actions, implementation dates and confirmed COVID-19 cases number from official reports of governmental institutions and official media in Algeria (Algerian Ministry of Health, 2020; APS, 2020).

### The preventive and control measures and strategy

The COVID-19 epidemic arrived in Algeria two months after the initial outbreak in Wuhan, China (WHO1, 2020), and a month later than the first case in Europe, January 24, 2020 (ECDC, 2020), which had given the stakeholders additional time to be prepared, so that, the national “media-planning” for surveillance, alert, health awareness and communication system has been activated since January, 2020, this was immediately when the WHO announced the spread of the SARS-CoV-2, subsequently, preventive measures were taken successively in accordance with international recommendations (WHO2, 2020) with a main objective, to reduce the effective contact rate, therefore, decrease the basic reproduction number. The preventive actions were mainly; social distancing, movement restriction, quarantine and lockdown.

As social distancing measures; On March 10, 2020, all sports, cultural and political meetings and fairs were cancelled, two days later, on March 12, 2020, all universities and educational institutions have been closed (APS, 2020), and on March 17, 2020, worship’s places were also closed (MARW, 2020). It was recommended to adopt good hygiene habits and to maintain a distance of at least one meter between people outside, however, it is important to mention that, until April 21, 2020, no instructions were given to incite ordinary people (not infected, not ill, not at risk and not taking care of a person at risk) to wear medical masks outside (Algerian Ministry of Health, 2020), this remains in line with WHO recommendations (WHO4, 2020).

Regarding movement restriction; On the international level, flights with China, Italy, Morocco and France have been restricted on February 3^rd^, 9^th^, 12^th^ and 15^th^, 2020, respectively, then, all land borders and air routes were suspended as of March 17, 2020, except aircraft carrying goods. Locally, all public transport means including rail traffic were suspended on March 22, 2020. In order to avoid cases importations and stem SARS-CoV-2 spread, a quarantine of 14 days in containment centres have been imposed on Algerian nationals repatriated since March 18, 2020 (APS, 2020).

Containment implementation; To efficiently detect, monitor and manage COVID-19 cases and to prevent further city-to-city transmission and avoid new regional outbreaks, more restrictive measures were taken on March 23, 2020, when the cumulative cases number reached a total of 234, and 125 cases (53%) in Blida only, thereby, more than one million inhabitants of Blida were placed under total lockdown and ban of movement to and from this region, furthermore, a partial containment was implemented and closure of unnecessary businesses in the capital city Algiers with the prohibition of any gathering of more than two people. Likewise, the partial containment was extended nationwide as of April 4, 2020, it was set up from 7 p.m. to 7 a.m. (12 hours) or from 3 p.m. to 7 a.m. (16 hour) depending on the epidemic situation in the region, and it is updated over time and regional outbreaks evolution. The response to these control measures mentioned above was estimated at 95% nationwide until April 6, 2020 (APS, 2020; Algerian Ministry of Health, 2020).

### The preventive strategy effectiveness assessment

The preventive measures and control actions were taken separately and gradually, so, we decided to evaluate the preventive strategy mentioned above in its entirety by considering March 23, 2020, as a strategy implementation’s date, from that day onwards, the main Algerian outbreak in Blida has been placed under full lockdown, and the other affected regions under partial containment.

We used the average basic reproduction number (R0) evolution as an indicator, so, we compared R0-before and R0-after the date assumed the preventive strategy would have effect, this represent 14 days after the strategy implementation, these 14 days are defined as the longer time COVID-19 symptoms are likely to appear after the last exposure according to WHO recommendations (WHO3, 2020), other study reported 11.5 days (97.5% CI 8.2-15,6) (Lauer, Grantz, & Bi, 2020). Thus, R0-before is estimated over the first 42 days of the epidemic (from February 25 to April 5, 2020), and R0-after is estimated over 14 days (from April 6 to April 19, 2020). The estimated R0-before and R0-after represent respectively the average R0 over the entire pre-implementation and post-implementation period of the preventive strategy, to this end, we calculated every day the R0 of the last 7 days, using equation 2, and we used a serial interval value SI=4.4 days (Chong, et al., 2020).

The COVID-19 cumulative cases divergence and projections based on R0-before and R0-after was done using the equation 1 of a predictive model established in a previous study (Hamidouche, 2020). In the end, we showed the epidemic projections on the basis of the results we found, this allowed us to predict the peak (Herd Immunity threshold) of the first wave of the epidemic using last week’s average R0 on equation 3.

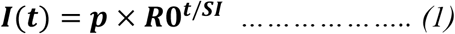

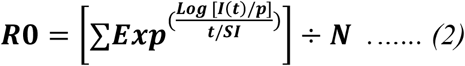

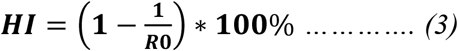

***R0:*** The average basic reproduction number

***I(t)*:** The incidence (cumulative cases number) at t time

***p*:** The baseline prevalence (in this study, **p** at 7 days before the estimation day)

***SI*:** The serial interval

***t*:** The time (in this study, 7 days)

***N*:** sample size

**HI:** Herd Immunity

### Statistical analysis and software

This study was carried out using Excel 2013 and STATA/IC 15 software. The bivariate analysis was conducted using Wilcoxon Mann Whitney test. We also used the Alg-COVID-19 Model established in a previous study (Hamidouche, 2020) to estimate the new projections and the epidemic’s peaks.

## Results

The outcomes of our observation and univariate analysis has revealed that R0 evolution since the beginning of the epidemic was very capricious, but, after the 40^th^ day of the epidemic that represent 12 day after mitigation actions implementation, the R0 evolution took a descending line in all three zones considered by the study (Fig.1.a.b.c). We also noticed that the second week’s average R0 were the highest across the whole country and in Blida at 4,26 (95% CI 2,65-5,88) and 3,28 (95% CI 2,28-4,27) respectively, but, outside of Blida, the highest value of R0 was in the 4^th^ week, it is equal to 3,2 (95% CI 2,44-3,95) after having been 1.09 (95% IC 0,87-1,28) the first week (Fig.2).

**Fig. 1.**
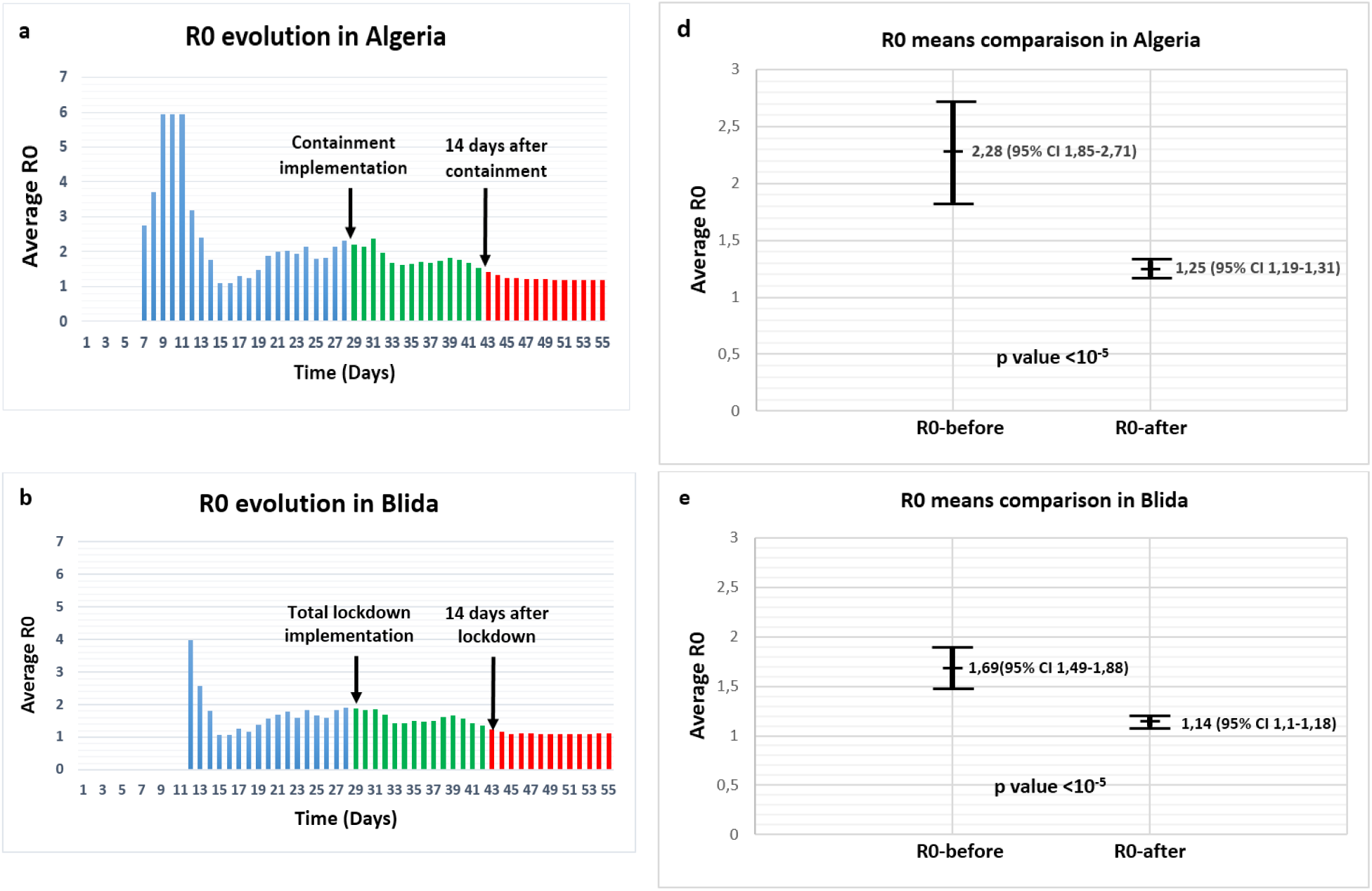
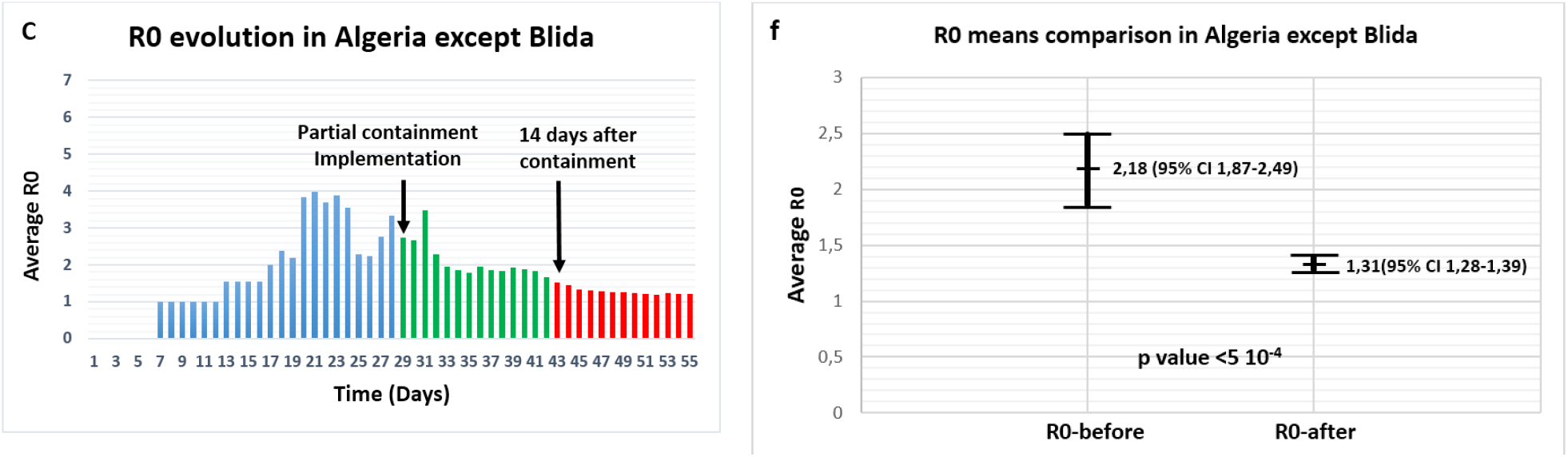
Univariate and bivariate analysis of R0 evolution before and after the preventive strategy implementation.

**Fig. 2.**
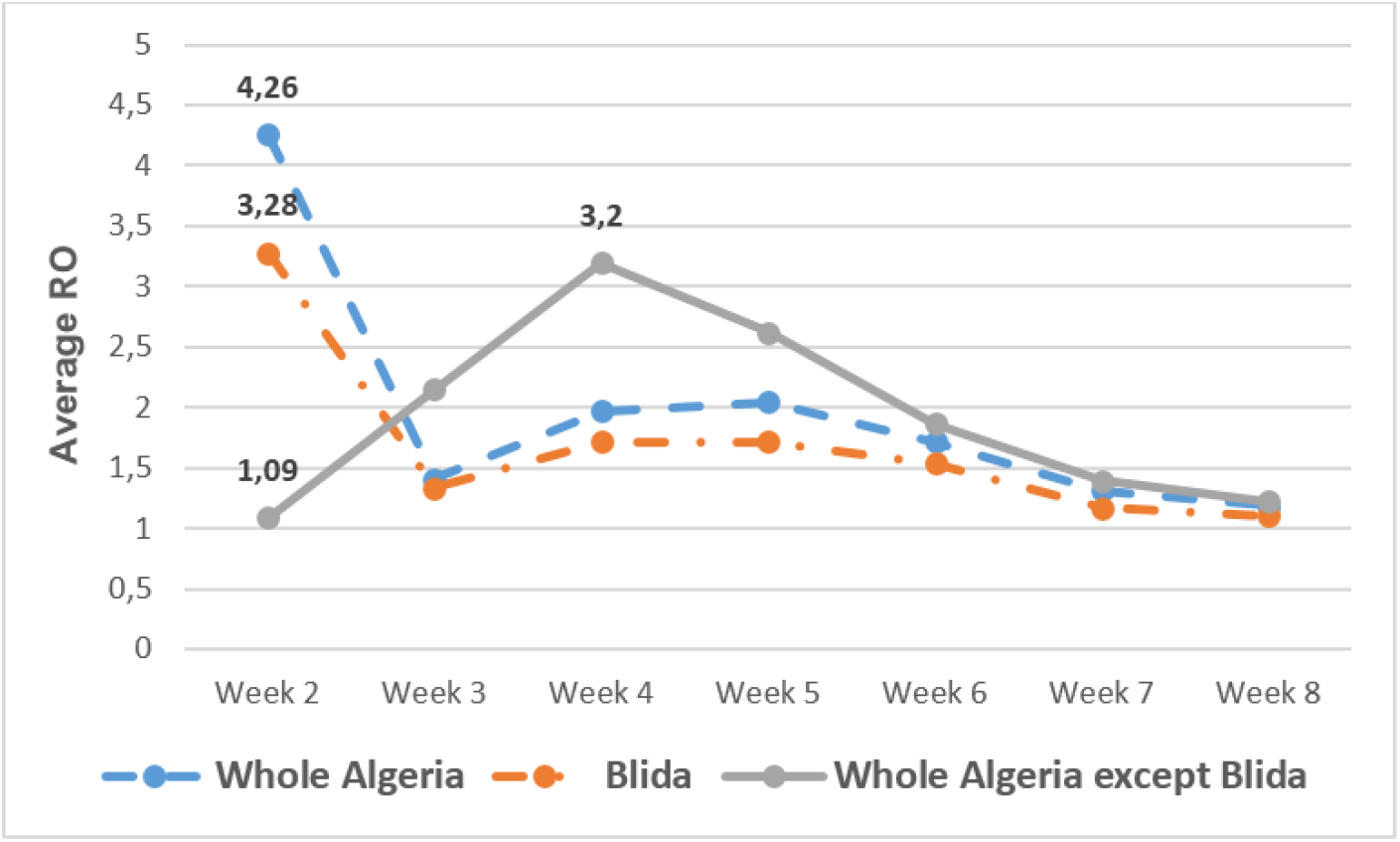
The weekly average R0 evolution in the three zones of the study.

The bivariate analysis conducted to compare the mean of R0-before (42 days from February 25 to April 5, 2020) and the mean of R0-after the preventive measures effect (14 days from April 6, to April 19, 2020) shows a statistically significant difference in the three areas considered by this study (Fig.1.d.e.f).

In Algeria entirely (Fig.1.d), the average R0 over the first 42 days of the epidemic was R0-befor=2,28 (95% CI 1,85-2,71) and R0 after preventive measures effect R0-after=1,25 (95% CI 1,19-1,31), the difference between the two means was significant with p value <10^−5^.

In Blida region (Fig.1.e.), the results are R0-befor=1,69 (95% CI 1,49-1,88) and R0-after=1,14 (95% CI 1,10-1,18), the p value was <10^−5^.

In the whole country except Blida (Fig.1.f.), the results show R0-befor=2,18 (95% CI 1,87-2,49) and R0-after=1,31 (95% CI 1,28-1,39), with p value <5 10^−4^.

In order to establish the divergence between the cumulative cases before and after the effect of the implementation of the control actions, we used the R0-before mentioned above, and the average R0 of the last week that we found in each zone, because, it represents the most updated values, so we used the following R0s; for the whole country: 1,19 (95% CI 1,18-1,20), for Blida: 1,10 (95% CI 1,09-1,11) and for whole country except the main outbreak of Blida: 1,22 (95% CI 1,20-1,24). We found that significant casualties due to COVID-19 have been avoided, and it would have occurred if the preventive measures had not been in place (Fig.3.a.b.c).

**Fig. 3.**
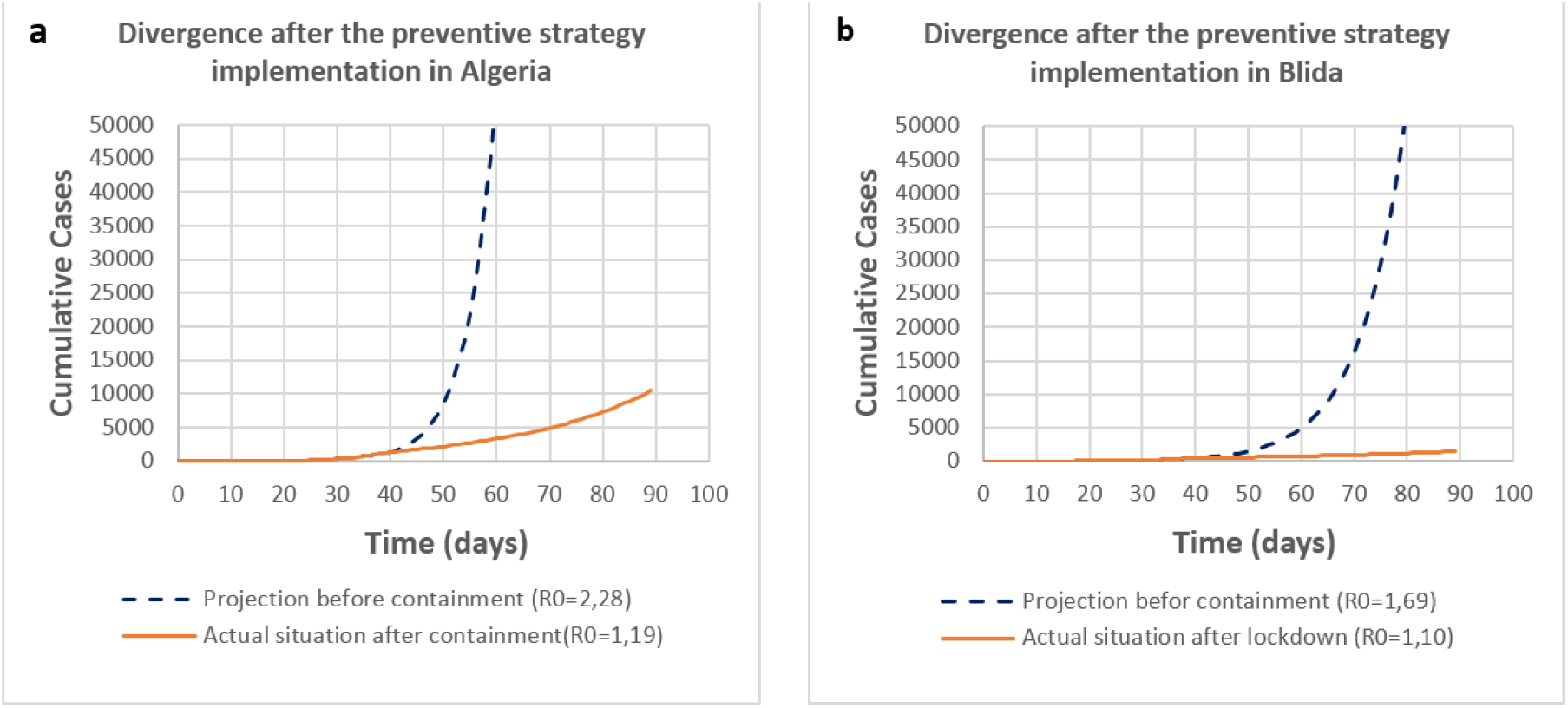
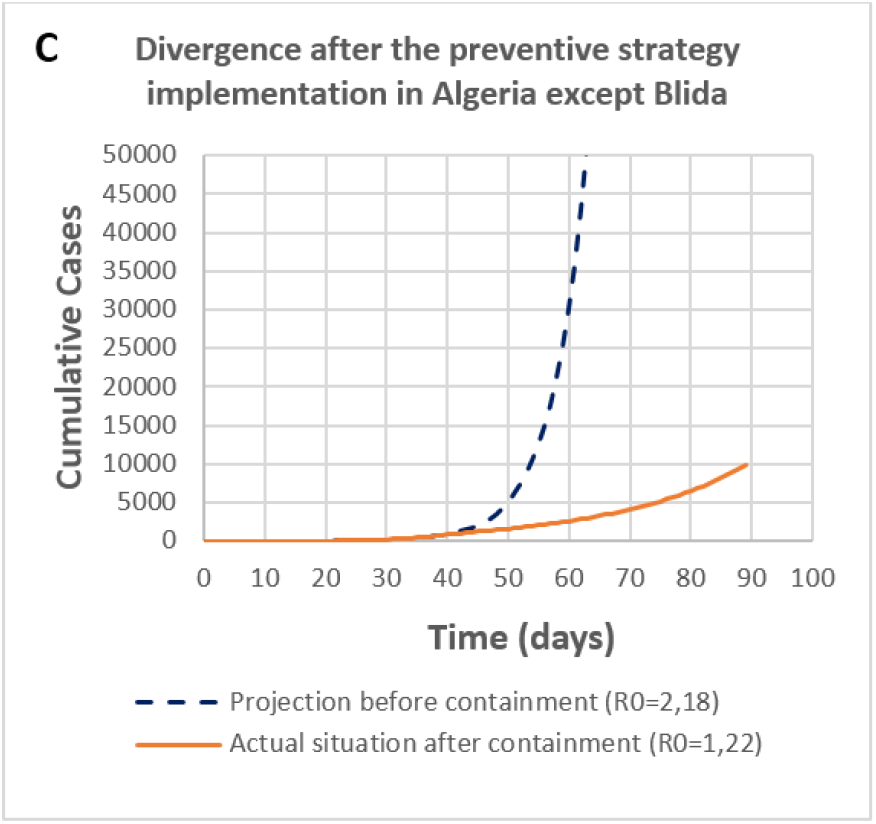
Divergence between actual cumulative cases and predictions before preventive measures implementations.

In Algeria entirely (Fig.3.a), according to our forecast results, the implemented preventive strategy has avoided the country 2993 COVID-19 cases (1914 instead of 4907 cases), and this only after the first week (from April 6, to April 12, 2020), over 14 days, 15613 infection has been avoided (from 6, to April 19, 2020), the actual cumulative cases were 2629 instead of 18242 predicted cases.

In Blida (Fig.3.b), on the first week, 495 infection has been avoided (1049 cases were predicted, 554 cases were reported), and 1747 prevented cases on two weeks (669 actual cases in lieu of 2416 infection).

In the whole country except Blida (Fig.3.c), the results show that we came out with 1633 fewer cases than what was expected to happen the first week (1360 instead of 2993 cases) and 7171 cases less if we add the second week (1960 rather than 10371 cases).

Based on the found R0 after the effect of the preventive strategy, we were able to predict the peak of the first wave of the epidemic, it represents the threshold of the Herd Immunity (HI), using the equation 3. If epidemic parameters are maintained like last week’s results, the peak in Algeria entirely will be achieved at HI=15,93%, in Blida at HI=9,57%, and in the whole country except Blida the peak will be reached at HI=18,37%. These HI thresholds that we have found predict an epidemic peak that will be achieved by the end of October to the beginning of November 2020 for the three zones.

## Discussion

Given the global impact of emerging and re-emerging infectious diseases, a number of methods have been developed to estimate and analyse the basic reproductive number R0, which is the hallmark of infectious diseases transmission (Blumberg & Lloyd-Smith, 2013), the most used approaches are based on four factor; i. the contact frequency, ii. the contact efficacy (both i & ii form the effective contact rate), in addition to; iii. the susceptible population size and iv. the infectious phase duration. However, these four parameters require time and numerous studies to be identified correctly, thus, in case of global emergencies such as the COVID-19 pandemic, we need quick results to assess the situation and the taken measures, especially in low and middle income countries where people in general are most affected.

Our assessment was mainly based on the evolution of R0, which was daily estimated using equation 2, taking as baseline prevalence 7 days before, we did not use the previous day prevalence to avoid variations due to postponement of cases declarations.

Monitoring of R0’s variations during the epidemic shows a remarkable progressive decrease over the last weeks (Fig.2), as mentioned in the results part, the highest R0 was recorded the second week at the national level and in Blida 4,26 (95% CI 2,65-5,88), and 3,28 (95% CI 2,28-4,27), respectively, as regards to outside Blida, the highest R0 equal 3.2 (95% CI 2,44-3,95) was reported two weeks later on the 4^th^ week. This is obviously due to the fact that the main outbreak occurred in Blida then spread outside.

On April 03, 2020, the effectiveness of the preventive actions has started to appear 12 days after their implementation (Fig.1.a.b.c), this observation is consistent with a previous study reported a symptom apparition duration of 11.5 days (97.5% CI 8.2-15.6) (Lauer, Grantz, & Bi, 2020), and also, in accordance with the 14-days containment recommended by the WHO for people likely to be infected (WHO3, 2020).

The statistically significant differences we found in the three areas after comparison between the two means R0-before and R0-after confirm the observations discussed in the paragraph above (Fig.1.d.e.f). Our results show that preventive measures that were implemented, whether total or partial containment, were generally effective, however, the total lockdown in Blida was more efficient in terms of decreasing the basic reproduction number to R0-after=1,14 (p-value<10^−5^) than the partial containment installed outside Blida, R0-after=1,3 (p-value<5 10^−4^). This situation can lead to two scenarios, the first scenario is controlling the epidemic in the next weeks by reducing R0 under 1, especially in Blida, the second scenario is displacement of the main outbreak from Blida to other cities with less strict preventive measures where the incidence is increasing, as like Algiers, Oran or Setif, with 460, 168 and 119 cumulative cases respectively as of April 21, 2020 (Algerian Ministry of Health, 2020), in the second scenario the herd immunity threshold and the peak dates that we have mentioned above need to be achieved in order to stop the first wave of the epidemic (Steven, et al., 2020). To avoid this, the preventive strategy can adopt a situation-updated plan, so that, the implementation of total containment can move to other cities or throughout the country in the coming weeks depending of epidemic’s displacement.

The projections we found show that thousands of COVID-19 cases have been avoided by the implementation of the previously mentioned preventive strategy, these numbers could be disputed by the underestimation of the actual cases numbers due to the low screening rate of COVID-19 in Algeria (6500 tests) (154 test/M population) until April 13, 2020, among them 1907(29%) were positive (only in the main laboratory) (El Watan, 2020), based on this fact, the difference between the actual cases and the projections that we found remains valid if we consider almost only symptomatic cases are reported, consequently, our projections represent almost only symptomatic cases.

We believe that the method we used to carry out the evaluation has given us a good overview of the effectiveness of the preventive strategy, and the basic reproductive number R0 is a precious variable to forecast the spreading of COVID-19, however, one of the limitation that we faced in this study is that, the R0’s estimating method that we used is based on the cumulative cases and not on the number of new cases reported daily, it was difficult to do so because of the large variation in the number of cases reported daily during the epidemic, therefore, this method can only detect the decrease in R0 up to 1 but not less. Furthermore, R0 is not the only factor to consider when estimating the burden of the epidemic and it is not necessarily the best indicator of the magnitude of the COVID-19 epidemic as it depends as mentioned above on the rate of symptomatic and asymptomatic cases and on testing capabilities. The most reliable parameters to assess the epidemic evolution remain first, the number of daily admissions to intensive care units, and secondly the emergency room admissions, and thirdly the number of hospitalizations. The observation of the daily number of deaths by COVID-19 is also good indicator if we take into account other bias such as age structure, quality of health service delivery and medical history of cases over time and places.

In order to better the control of the epidemic, we should decrease the basic reproduction number, no drugs or vaccines have proven their efficacy yet, so, to diminish the effective contact rate we need to act on its two components; the contact frequency and the contact efficacy. The preventive strategy in Algeria was mainly contact frequency reduction by containment and quarantine, this will reveal new regional outbreaks, and will help identify and isolate infected people and their surroundings with screening tests if available, on the other side, it is possible to act on contact efficacy by adding to good hygiene habits, an instruction to wear medical mask by ordinary population outside, this measure has been reported to be effective in some countries (Kar, Tai, & Chi, 2020), especially that pre-symptomatic transmission has been reported; exposure in these cases occurred 1 to 3 days before the source patient developed symptoms (Wei, et al., 2020), and pre-symptomatic transmission may contribute to 48% and 62% of transmissions (Ganyani, et al., 2020), however, this action may encounter some supply difficulties due to the lack of medical masks on the market.

The Herd Immunity (HI) thresholds mentioned in the results section (whole country HI=15,93%, Blida HI=9,57%, and in the whole Algeria except Blida HI=18,37%) are the peaks of first wave of the epidemic, these are insufficient levels to avoid a second epidemic wave if the preventive measures will be lifted after the date indicated, regardless the control actions of any country, the epidemic will stop only if the final HI threshold based on the COVID-19 initial R0 without control actions is achieved or maintain R0 under 1 for a considerable duration with a sever lockdown, but, this seem to be hard to do. The initial basic reproduction number R0 in Algeria was 2.28, for this value, at least 56% of people with protective immunity would be needed to be sufficient to avoid a second wave, because the actual COVID-19 seroprevalence in Algeria is unknown and difficult to estimate, we cannot predict for the time being the end of the epidemic.

To conclude, this study shows that the implementation of the prevention strategy as a whole is effective in Algeria and it succeeded to maintain the epidemic growing at a very slow pace. However, it is good to keep in mind that a new way of life based on good hygiene habits and social distancing must therefore be in place and adopted by the population for at least several months, otherwise the demand for health care will exceed the availability.

## Data Availability

all data is vailable.

